# Role of high-dose exposure in transmission hot zones as a driver of SARS-CoV2 dynamics

**DOI:** 10.1101/2020.10.07.20208231

**Authors:** Dominik Wodarz, Natalia L. Komarova, Luis M. Schang

## Abstract

Epidemiological data on the spread of SARS-CoV-2 in the absence and presence of various non-pharmaceutical interventions indicate that the virus is not transmitted uniformly in the population. Transmission tends to be more effective in select settings that involve exposure to relatively high viral dose, such as in crowded indoor settings, assisted living facilities, prisons, or food processing plants. To explore the effect on infection dynamics, we describe a new mathematical model where transmission can occur (i) in the community at large, characterized by low dose exposure and mostly mild disease, and (ii) in so called transmission hot zones, characterized by high dose exposure that can be associated with more severe disease. Interestingly, we find that successful infection spread can hinge upon high-dose hot zone transmission, yet the majority of infections are predicted to occur in the community at large with mild disease. This gives rise to the prediction that targeted interventions that specifically reduce virus transmission in the hot zones (but not in the community at large) have the potential to suppress overall infection spread, including in the community at large. The model can further reconcile seemingly contradicting epidemiological observations. While in some locations like California, strict stay-home orders failed to significantly reduce infection prevalence, in other locations, such as New York and several European countries, stay-home orders lead to a pronounced fall in infection levels, which remained suppressed for some months after re-opening of society. Differences in hot zone transmission levels during and after social distancing interventions can account for these diverging infection patterns. These modeling results warrant further epidemiological investigations into the role of high dose hot zone transmission for the maintenance of SARS-CoV-2 spread.

## Introduction

As the United States and other countries around the world have witnessed the first wave of SARS-CoV-2 spread and the associated morbidity and mortality, it is clear that a balance between non-pharmaceutical interventions that limit the public health burden on the one hand, and a continued functioning of society that limits economic damage on the other, is required until other means to combat the pandemic are available (such as effective vaccines). Non-pharmacological management interventions appear to be the most appropriate approach in this context. Mathematical models have been used to characterize the dynamics of SARS CoV-2 and predict potential numbers of COVID-19 cases [1-7], which has resulted in the estimation of the basic reproduction number [1, 8], a better understanding of expected transmission dynamics in the absence and presence of non-pharmaceutical interventions [9-16], and in the critical effect of age structure on disease dynamics [11, 17], among many other contributions. Some of these models have been extremely useful for predicting and quantifying the demands on health care resources.

At the same time, it is becoming clear that the spread of SARS-CoV-2 is characterized by unique aspects that have so far not been taken into account by epidemiological models and that might be crucial for predicting how the virus and the disease may spread, depending on the degree to which the society and economy is open. The data on infection spread during various phases of lockdown and re-openings in the US indicate that the virus does not spread uniformly in the population, but that some settings contribute more to virus transmission that others[18]. In California, initial stay-home orders in March prevented continued virus spread, but failed to drive the effective reproduction number significantly below one and to reduce the existing infection prevalence. After a significant surge occurred following a wide-spread re-opening of society and businesses, more targeted interventions were implmented, under which business activities that involved large indoor gatherings (such as bars, indoor seating in restaurants, or indoor hair salons) were restricted. This resulted in a significant reduction in virus prevalence to levels that are similar to those during the stay-home orders, despite the fact that many other aspects of society, such as outdoor business operations or activities in the community at large remained mostly unrestricted. An increased focus on mask wearing, which minimizes viral shedding and thus environment load, might have also contributed. These trends indicate that similar levels of infection control can be achieved whether society as a whole is totally shut down, or more select aspects of society are preferentially restricted. Similar patterns have occureed in other states, or countries (e.g. most of Europe). While strict shut-downs of society have brought infection levels under control in New York and in countries such as Germany, the UK, or Denmark, the infection then remained controlled for several months following a reopening of society. During those times, although most restrictions were lifted for the community at large, certain restrictions remained in place. Those included infection control measures in critical places such as hospitals and long-term assisted living facilities (which minimize infection of the elderly), as well as avoiding large indoor gatherings. Indeed, it appears that local outbreaks during this time may have been driven by infection spread in select settings. Examples include food processing plants [19], assisted living facilities [20], or hospitals [21].

These observations indicate that overall viral transmission appears to occur mostly in the context of select settings, in which people are exposed to relatively high doses of the virus, and less so in the community at large, where viral doses tend to be lower. Viral dose upon exposure can influences the chances of developing a productive infection and can impact the severity of disease. Viral infectious dose has drastic consequences for SARS and MERS infections [22, 23], and for pathogenesis of SARS COV-2 in animal models [24, 25]. Recent evidence strongly suggests that reducing viral infection load by using facemasks has a pronounced effect on the outcome of human infections [26]. Considering this evidence, one can presume that situations that lead to exposure to higher viral doses may well drive a substantial portion of SARS CoV-2 spread. These situations can be collectively referred to as “transmission hot zones”, and comprise physical locations such as long term assisted living facilities (which also tend to house older, more susceptible, people), prisons, food processing plants, and bars, among others, or transient situations of large or periodic recurrent gatherings [27]. Understanding the principles of hot zone-driven infection spread requires the incorporation of these assumptions into mathematical models, in particular the assumption that the rate of disease transmission, as well as disease severity, depend on exposure dose. Hot zone transmission is defined by settings in which people are exposed to a higher virus dose than in the community at large.

Here, such a mathematical model is constructed and analyzed with special focus on the basic reproduction number. We find that, somewhat counterintuitively, successful infection spread can hinge upon hot zone transmissions that promote severe infections, yet at the same time the majority of the SARS-CoV-2 infections are mild and occur in the community at large. The model further predicts that in such cases, targeted interventions that limit virus spread in hot zones can result in the long-term suppression of infection levels in the community at large, even if non-pharmaceutical interventions in the community are relaxed to some extent. According to our mathematical model, these dynamics are a direct consequence of the assumed viral dose-dependency, which might thus warrant further attention from a clinical and epidemiological perspective.

## Results

### The mathematical modeling framework

We consider a mathematical model that distinguishes between patients with mild or asymptomatic infection and those with severe SARS CoV-2 infection, including symptomatic but ambulatory COVID-19 patients. We further assume that a higher infectious dose promotes development of more severe outcomes, as has been documented with SARS, MERS, and even SARS CoV-2 [22-24, 28]. In particular, the model couples viral load to the setting in which transmission takes place. Hence, we distinguish between two basic types of transmission varying in the viral load to which susceptible individuals are exposed (Fig 1). The first type of transmission occurs in the “community-at-large”. A characteristic of this environment is that people are exposed to relatively low viral loads for short times, and that disease tends to be mild. This can include streets and other outdoor areas, as well as indoor locations where it is unlikely that several infected individuals converge periodically for long periods, and where human density is low and contacts between individuals are short and occasional. The second type of transmission is in what we call “hot zones”. These are characterized by exposure to high viral loads and by a higher chance of severe disease. This can result from exposure to multiple infected individuals, exposure to individuals shedding high viral levels in locations with poor ventilation, or from an increase in viral load over time through silent amplification rounds of infection[29]. The model is based on the general SIR framework [30, 31]. The corresponding ordinary differential equations are given as follows:

**Figure 1.**
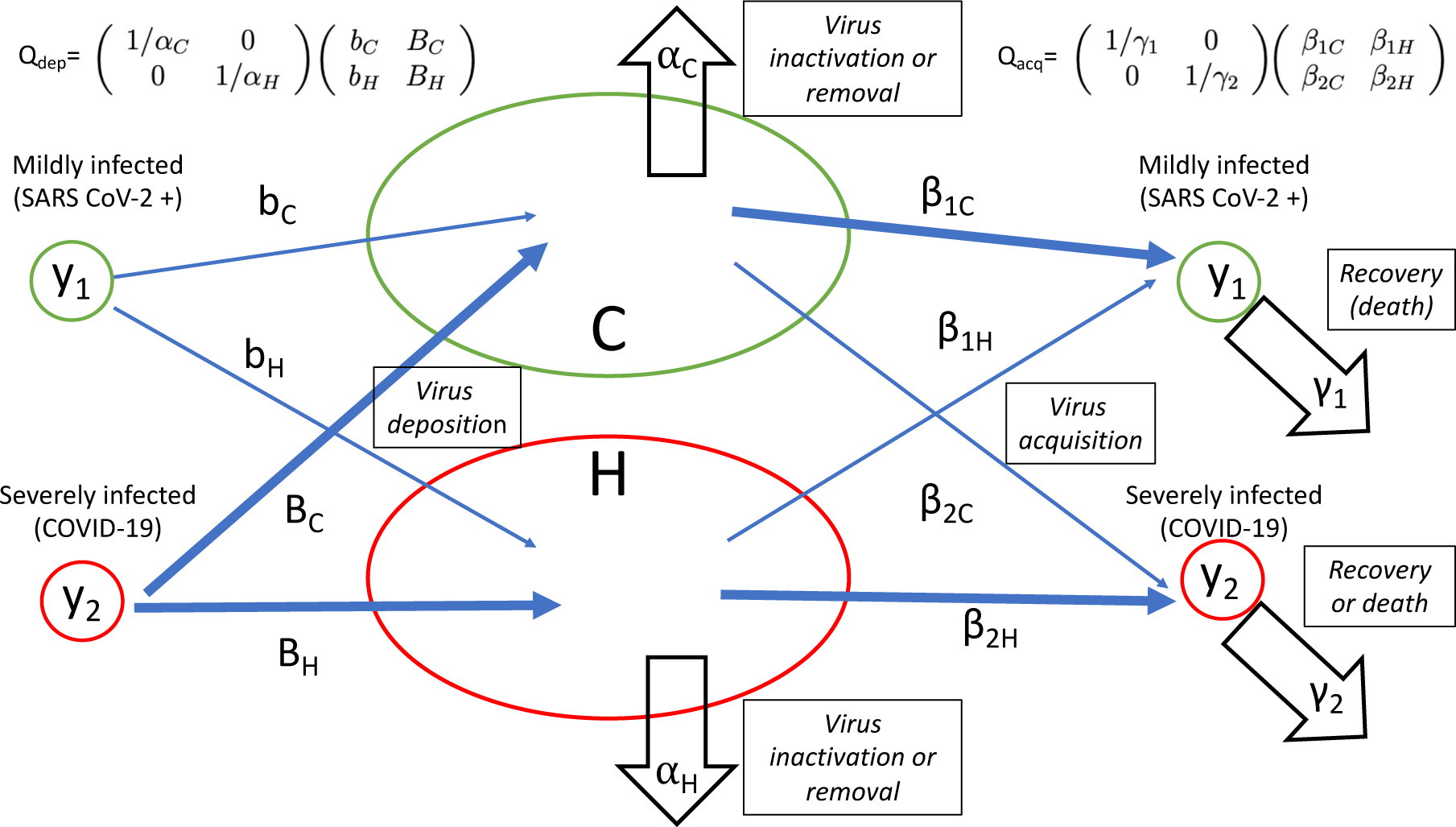
A schematic showing the model structure and its parameters. Infection deposition and acquisition matrices, Q_dep_ and Q_acq_ are defined, see Supplement for details. In the model, transmission can result in two types of infected populations: asymptomatic/mild SARS CoV-2 infection (denoted by y_1_) and severely SARS CoV-2 infected, including ambulatory symptomatic COVID-19 patients (denoted by y_2_). It is assumed that severe COVID-19 is promoted by exposure to a higher viral load. Hence, exposure of susceptible individuals to virus in the community at large compartment (C) results mostly in mild infection with a probability β_1C_, and less frequently in severe infection with a probability β_2C_, where β_1C_> β_2C_ because local viral load is assumed to be relatively low in the C compartment. Exposure in the hot zone compartment (H) results in mild infection with a probability β_1H_, and in more frequent severe COVID-19 with a rate β_2H_, where β_1H_<β_2H_, because virus load is assumed to be higher in the hot zone. Mildly infected individuals are assumed to deposit virus in the C and H compartments with rates b_C_ and b_H_, respectively. Severely SARS CoV-2 infected, including symptomatic but ambulatory, individuals are assumed to deposit virus in those compartments at rates B_C_ and B_H_, respectively. Finally, virus decays in the two locations at rates α_C_ and α_H_, and mildly and severely infected individuals cease to be infectious (because of recovery or death) with rates γ_1_ and γ_2_.

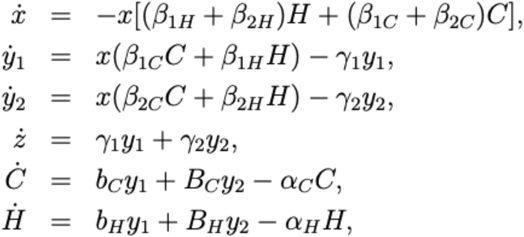

where x denotes the population of susceptible individuals, y_1_ and y_2_ denote the populations of mildly and severely infected individuals, respectively, and z represents the population of removed infecteds (recovered and dead). Further, C and H represent environmental viral load in the community at large and in the hot zones, respectively. The processes underlying the model are further explained in Fig 1, where all the parameters are defined. The following sections discuss results arising from this model, and further mathematical details are provided in the Supplementary Materials.

### The basic reproduction number and maintenance of infection spread

We use this model to calculate the basic reproduction number of the infection, R_0_, as well as the effective reproduction number, R. We start by defining two matrices, a virus deposition matrix, Q_dep_, and a virus acquisition matrix, Q_acq_. The former matrix is given by

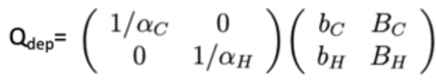

and depends on the rates at which mildly and severely infected individuals deposit the virus both in hot zones and the community, and also on the virus life-span in each environment. The latter matrix is defined as

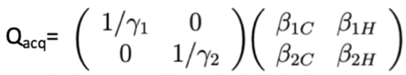

and depends on the infectivity coefficients for mildly and severely infected individuals in both environments, as well as the expected disease duration for mild and severely infected patients. To determine the reproductive number (including the basic reproductive number) of the infection, we form a product of the acquisition and deposition matrices,

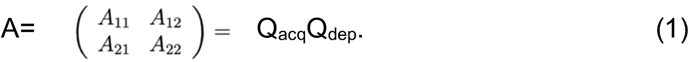

The elements of this matrix, A_ij_, are combinations of all of the rates, see Fig 1 and the Supplement. These four quantities have a clear meaning, as they express the intensity of infection spread via four pathways: A_11_ is the probability to become mildly infected as a result of another mildly infected individual depositing virus in a C or an H location; A_12_ is the probability to become mildly infected as a result of a severely infected individual depositing virus in a C or an H location, etc.

Using the matrix A, the basic reproductive number of the infection can be expressed concisely as

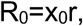

where x is the initial number of susceptible individuals and *r* is the larger of the two eigenvalues of the matrix A. Similarly, the effective reproductive number, R, is calculated as R=xr, where x is the current number of susceptible individuals. The quantity r depends on the model coefficients, and will be affected e.g. by social distancing measures.

Depending on the parameters, the virus may spread faster through some pathways than others. For example, if A_11_ in equation (1) is significantly larger than the other matrix elements, then we simply have R≈xA_11_, that is, infection spread mostly occurs from mildly SARS CoV-2 infected individuals to result in more mildly infected individuals, and the kinetic parameters associated with mild infection define the R value of the whole system:

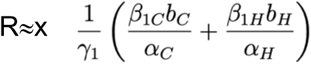

On the other hand, if the element A_22_ is significantly larger than the rest of the matrix elements, we have R≈xA_22_, and it is COVID-19 severely infected individuals that maintain the epidemic:

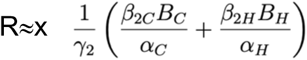

The off-diagonal elements A_12_ and A_21_ define the contribution of one group of infected to the expansion of the other group. The relative size of these pathways influences the population sizes of y_1_ and y_2_. For example, A_12_>A_21_ tends to increase the population of the mildly infected. The opposite inequality results in the boosting of the y_2_ population, see Supplement for a more precise statement.

Depending on the relative values of the matrix elements A_ij,_ this model can give rise to a variety of different dynamics and outcomes. To understand this better, it is instructive to first consider extreme cases that bracket all possible outcomes.

- (Ia): Maintenance of the epidemic depends on the mildly infected individuals, y_1_, transmitting virus through the C (or H) compartment and giving rise to mostly more mildly infected individuals. Most infected individuals have mild disease, whereas given subsets develop serious COVID-19.
- (Ib): Maintenance of the epidemic again relies on transmission by y_1_ individuals creating more y_1_-infected people, but most infected individuals are y_2_ and have severe disease.
- (IIa): Maintenance of the infection depends on severely infected individuals, y_2_, transmitting the infection (through C or H). The majority of the infected individuals, however, are y_1_ and have mild disease, whereas given subsets develop serious COVID-19.
- (IIb): Maintenance of the infection again depends on severely infected individuals, y_2_, but most of the infected individuals have severe disease.

While not all of these cases are realistic, an important and novel point emerges from this analysis: It is possible that the group of infected individuals that is responsible for maintaining the epidemic (i.e. for keeping R_0_>1) comprises only a small subset of the infected people. For example, in case (IIa), severely infected individuals that transmit the virus to generate new patients with severe infection, including serious COVID-19 (via hot-zone transmission) is critical for keeping R_0_>1. At the same time, however, the majority of the individuals is mildly infected, y_1_, see Fig 2(b). This gives rise to the possibility that the targeting of a relatively small fraction of infected individuals in hot zones through specific interventions could curb overall SARS CoV-2 prevalence in the community at large.

**Figure 2.**
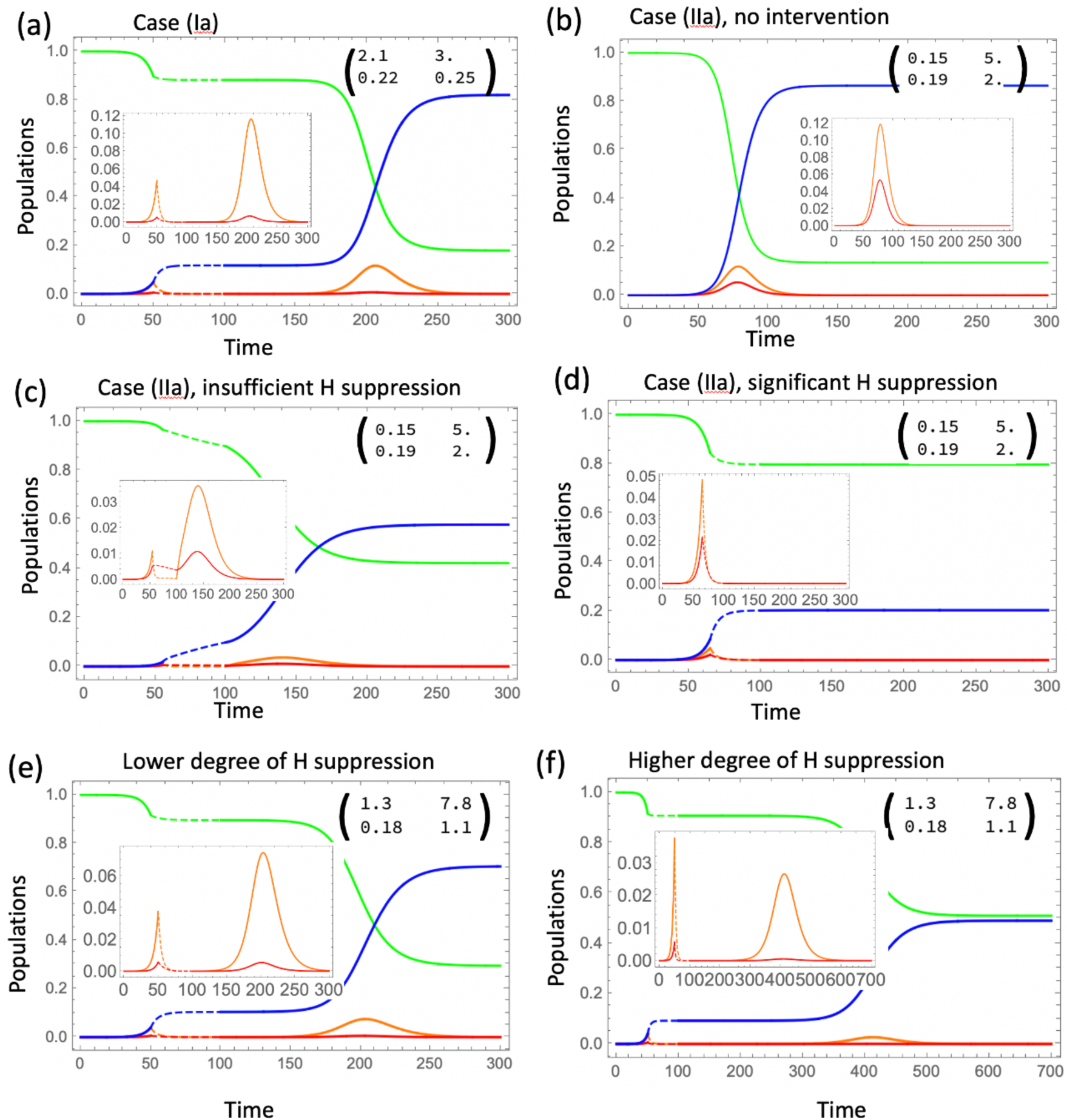
Simulated epidemic dynamics in different cases. Four populations are shown as functions of time: susceptible (green), mildly SARS CoV-2 infected (orange), severely COVID-19 infected (red), and recovered/dead (blue). Insets show detail in the dynamics of infecteds, and the matrix elements A_ij_. Intervention is shown by dashed lines. It is assumed that during intervention, both H and C channels are suppressed, and afterwards channel C is restored to its full capacity. (a) Case (Ia) (see text), where the spread of infection is mostly through mildly infected who comprise the majority; opening up leads to a second wave of infection. (b-d) Case (IIa), where the spread of infection is mostly through severely infected although the majority are still mildly infected. In (b) no intervention is implemented; the effects of social distancing are shown in (c) (insufficient H suppression, a second wave is predicted) and (d) (significant H suppression, no second wave). (e,f) Both C and H channels contribute about equally to infection spread; opening up the C channel results in a second wave, but a higher degree of H suppression leads to a smaller and more delayed second wave. In all simulations, R_0_=2.4. Other parameters are given in Fig S2 and S4.

### Simulating non-pharmaceutical interventions and re-openings

Some valuable insights can be gained by simulating the implementation of non-pharmaceutical interventions and their relaxation. At time t_1_, social distancing is initiated by parameter changes that promote a reduction in the reproduction number, e.g. by reducing virus deposition rates (b_C_, b_H_, B_C_, and B_H_) and infection rates (β_1C_, β_2H_, β_1H_ and β_2C_), or by increasing virus removal rates (α_C_,α_H_) and patient removal rates through quarantining (γ_1_,γ_2_) (Fig 1). At time t_2_, re-opening is simulated by reverting most parameters back to their original values, with the exception of select parameters connected to either C or H transmission.

We focus on realistic cases, where mild infections predominate. For non-pharmaceutical interventions, we simulate the stay home orders that were implemented around March-April 2020, where it is assumed that virus transmission in the community at large is significantly suppressed, but that hot zone transmission may or may not continue to operate. When simulating the opening of society, different assumptions are made about the extent to which virus transmission resumes in the community at large and in the hot zones.

#### Community at large transmission alone maintains infection spread

First, we assume that maintenance of the epidemic depends only on community spread and that hot zone transmission contributes little (case (Ia) above). This corresponds to a scenario in many previously published COVID-19 models, e.g. [13]. Under this assumption, we observe that the suppression of virus transmission in the community at large leads to a marked reduction in infection prevalence. Upon re-opening, a pronounced second wave of infection ensues until a vast majority of individuals in the population have been affected (Fig 2a).

#### Hot zone transmission alone maintains infection spread

Next, we assume that maintenance of the epidemic relies on hot zone transmission and that community transmission contributes little (case (IIa) above). We distinguish between two scenarios. First we assume that during stay-home orders, hot-zone transmission continues to occur. We then assume that hot-zone transmission is also suppressed.

i. Assume that high viral load hot zone transmission remains elevated in certain pockets during the interventions, such that the overall reproduction number is slightly larger than one. In this case, the infection continues to spread slowly during social distancing, and the majority of the infections are predicted to be mild. Under this scenario, infection prevalence is predicted to not decline during the lockdown, similar to the dynamics observed in California. Upon reopening, a renewed and accelerated spread is always predicted to occur. Because the reproduction number continues to be larger than one during the intervention period, it can only increase further during relaxation of the interventions (Fig 2c).
ii. If hot zone transmission is suppressed during the stay-home interventions, virus prevalence markedly declines during the intervention period (Fig 2d). If re-opening only leads to resumption of virus transmission in the community at large, and hot zone transmission remains suppressed, no second wave is predicted to occur because the reproduction number remains below one (Fig 2d). If, however, hot zone transmission increases after reopening (due to re-activation of previous hot zones or generation of new ones), the reproduction number can increase beyond one, and a second wave happens in the model (not shown). This scenario might correspond to dynamics observed in New York and several European countries: Stay-home interventions resulted in significant suppression of the infection, and this level of suppression was maintained for 2-3 months even after the initial re-opening of society, presumably because of the continued suppression of hot zones. As the infection levels started to rebound e.g. in Germany, this was associated with outbreaks in hot zones, such as food processing plants[19].

#### Both hot zones and the community at large can maintain virus spread

Here, we assume a more balanced contribution of hot zones and the community at large: either pathway alone can lead to sufficient transmission such that R>1, but both transmission pathways are needed for the virus to achieve its full spread potential. Different outcomes are possible depending on the particulars. For stay-home interventions to lead to a marked reduction of infection levels, transmission reduction has to occur both in the community at large and in the hot zones. If hot zone transmission is not significantly affected by the intervention, infection spread will be slowed, but no decline will occur (not shown). In any case, reopening is likely to lead to a second wave, even if virus transmission only resumes in the community at large. The magnitude and timing of the second wave depends on social distancing and reopening parameters. For example, if hot zone transmission is suppressed to a larger degree, the second wave will be characterized by a slower growth rate and a lower size (Fig 2(f) compared to 2(e), note the different range of the horizontal axes and the lower final epidemic size).

### Further Complexities

Additional insights can be obtained by incorporating further complexities that might better characterize the SARS CoV-2/COVID-19 pandemic. A mathematical model that includes these assumptions is presented in the Supplementary Materials. In the example of Fig 3 we assume that once the virus load in the hot zones rises to high levels, it is likely that the rate of virus transmission saturates rather than increases in an unlimited way. Further, we assume that severity of infection is not only transmission zone-dependent, but that a higher viral load in both C and H locations results in a higher chance of severe infection. The model now predicts that the fraction of severe infections rises as total infection levels increase, but that it then declines post-peak (Fig 3(b)). This might correspond to the observations of increased disease severity as the epidemic expands, and then reduced disease severity as the outbreak declines, which has been observed in Italy [32] and Sweden [33], among other locations. Importantly, the simulations in Figure 3 show that during the initial stages of spread, mild SARS CoV-2 infections predominate, but once the infection has spread beyond a certain threshold, the total viral load in both hot zones and the community at large increases to the point that severe COVID-19 cases predominate. This would translate into a health crisis with overload of the health systems and into high levels of mortality even if health care resources did not become limiting. According to this model, it is critical to implement interventions sufficiently early rather than only once increased mortality becomes apparent (which occurs with an additional delay).

**Figure 3.**
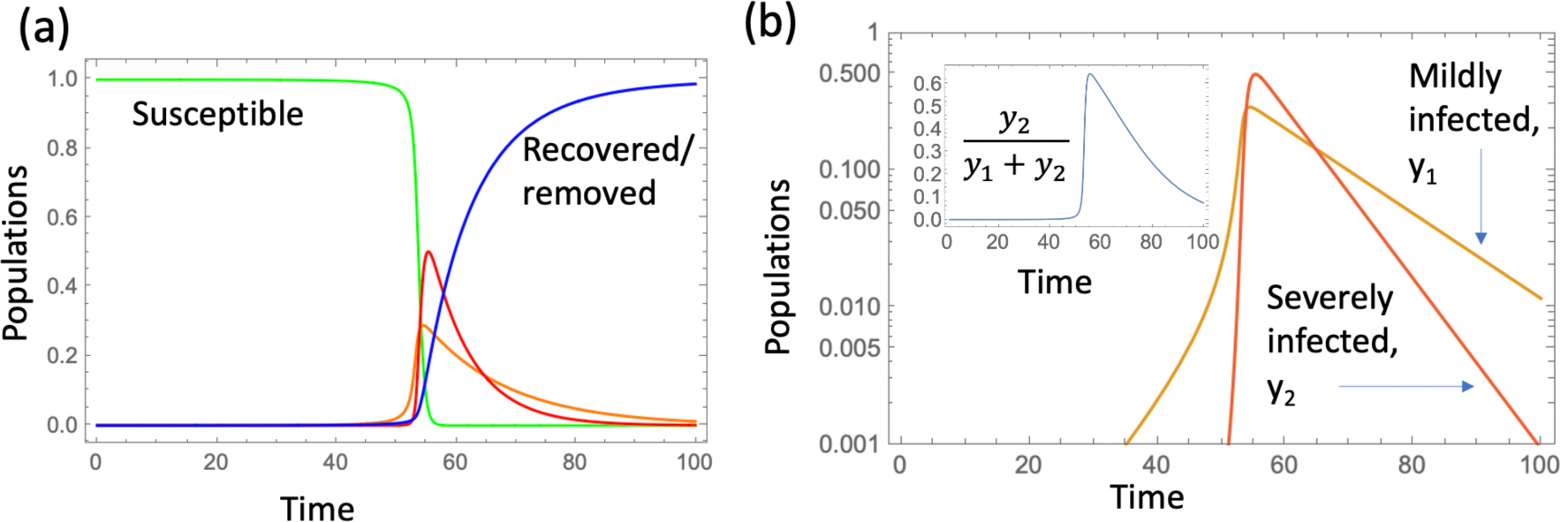
More realistic scenarios of infection spread: included is a saturation term in the C and H channels, and the probability of becoming severely infected through channels C and H depends on the infection level, resulting in a change in the proportion of severely infected. The epidemic dynamics are shown in (a) and the fraction y_2_/(y_1_+y_2_) in the inset in (b). For all parameter values, see figure S4.

## Discussion

We have presented and analyzed a new epidemiological model in which infection severity depends on the virus dose received during transmission, assuming that exposure to higher virus loads is more likely to occur under certain conditions, which we refer to as transmission hot zones. These high viral loads can be reached by silent, undetected amplification cycles before the first clinical cases are detected [29]. One important insight was that a relatively small population of severely infected patients can be responsible for driving infection through hot zone transmission, even though the majority of infections are mild and acquired in the community at large. A logical consequence of this is that even though the majority of infections are found in the community at large, interventions that specifically target the hot zones can be very effective. They can reduce the viral reproduction number, inducing a decline of the overall infection prevalence, including that in the community at large. Hence, effective virus suppression in hot zones might allow to maintain a relatively strong degree of overall infection control with fewer restrictions in the community at large, although critical interventions in the community at large will most likely also be required.

The model can further explain some seemingly contradictory observations. For example, in California, stay-home orders halted further infection spread, but a significant decline of infection levels was not observed during this phase. According to our model, an effective reproduction number that is larger than one can be obtained during interventions by continued viral transmission in yet unidientified hot zones, even though infection spread in the community at large is strongly inhibited due to the stay home orders. In agreement with this hypothesis, a large fraction of the recorded infections and deaths occurred in nursing homes and assisted living facilities in California during the time period when the stay-home order was in place [34]. On the other hand, in other locations, such as New York and several European countries, lockdowns were more effective at driving infection numbers down, and when society subsequently opened up to a certain extent, infection levels appeared to remain controlled for several months. According to our model, this would be explained by more efficient suppression of viral transmission in the hot zones during lockdown, and continued hot zone suppression after the initial reopening phase (e.g. by protecting assisted living facilities and hospitals, and by avoiding larger indoor gatherings in bars etc.). The model further makes the prediction that renewed hot zone transmissions would be able to trigger a second wave. This is again consistent with observations. For example, in Europe, the initial growth in case numbers during the emerging second waves seemed to be associated with hot zones, such as food processing plants [19].

This epidemiological model also provides a motivation to act early and decisively to prevent the amplification of the viral loads and to prevent potential transmission hot zones with severe disease from forming. Facemasks might be crucial in this respect, because they reduce exposure dose. Facemasks can thus turn a potential hot zone that drives infection spread into a lower dose transmission environment, which does not have the same ability to maintain overall infection spread.

Past coronavirus outbreaks have been characterized by large numbers of infections resulting from superspreading events; the corollary to this finding is that most individuals infected with these viruses did not transmit the virus efficiently [18, 35, 36]. The “hot zone” transmission framework proposed here is broadly consistent with these findings: hot zones are characterized by the exposure of susceptible individuals to a relatively high virus dose, which may lead to more severe disease. Large gatherings, including several infected individuals shedding virus at the same time, can provide the high dose exposures, as could amplifications of the viral load by the repeated visits to a location by the same people. While early virus exposure from one infected person would likely constitute a low dose exposure and result in mild infection in several individuals, these individuals would then all shed virus, thus increasing the viral dose with which others are infected. Hence, amplification through asymptomatic or mild cases will eventually result in exposure to a higher virus dose and in a higher chance of severe disease in the newly infected people, thus generating a hot zone environment. Such characteristics could exist in crowded/ poorly ventilated office work spaces, schools, prisons, places of worship, long term care/assisted living communities, and health care/hospital settings, as well as in multigenerational households, which have all been identified as transmission clusters in various countries [18]. Long term care facilities and hospitals might be especially prone to this effect, which is consistent with the large percentage of deaths in long term care facilities. People with pre-existing health conditions regularly visit hospitals, which could start such an amplification cycle anywhere in a hospital before the presence of the virus in the location is known.

These notions are consistent with data from Italy, where at least some hospitals were identified as a major contributor to early COVID-19 spread, like in Bergamo [37]. Similarly, in the United States, an infection cluster has been recently identified in a hospital in Boston [38]. These notions are further consistent with the many analyses of the disproportionately high mortality of elderly people residing in long term care living facilities [39, 40] rather than at individual homes across the world [41], and with the disproportionate impact on minority communities in America [42], who may have been required to physically go to work during the outbreak due to their employment in “essential services” like transportation and food preparation. A number of Asian countries implemented strong protection measures in the health care system before the first COVID-19 cases were identified, due to previous experiences with SARS and MERS. Countries such as South Korea performed intensive testing, resulting in early identification of infected individuals, which were thus removed from both transmission chains and any potential amplification circles [43]. Such strategies limit the seeding of transmission hot zones and prevent the initial rounds of viral load amplification, which might have contributed to the relatively lighter disease burdens documented there, despite somewhat less strict social distancing [44].

This model can be refined by including risk factors, social networks and other complexities, which all are likely to be critical for any practical predictive use. For example, we assumed in our model that disease severity was associated with the dose of virus exposure. Risk factors (importantly age and comorbidities), however, are another important determinant of disease severity, and incorporation of these additional details will refine the accuracy of the model. Moreover, model predictions depend on assumptions, which need to be tested with epidemiological, clinical, and virology data. The strength of modeling, however, is to identify potential key drivers of the pandemic, which we would not be aware of otherwise, thus directing the required epidemiological, clinical, and virology investigations. The finding that severe infection transmission through high viral load exposure in hot zones might be an important driver of SARS CoV-2 spread, even as mild disease cases predominate, could allow us to improve the outcome of reopening society through targeted interventions.

## Materials and Methods

We have modeled the spread of infection by using ordinary differential equations (ODEs) of SIR type, where we distinguished between patients with mild or almost asymptomatic infection and those with severe SARS CoV-2 infection. Transmission happened through two different “channels”. Several extensions of the model are also considered including a model with saturation in the infection term. The precise formulations and complete analysis of the ODEs are presented in the Supplementary Materials.

## Data Availability

The manuscript is concerned with mathematical models and does not contain data.

## Acknowledgments

All authors wish to thank Dr. Nathaniel Hupert, MD, MPH (Weill Cornell) for most helpful editions, and LMS also for many stimulating discussions. This work was funded in part by NSF DMS 1662146/1662096 (NK & DW). LMS is funded by NIH AI153396 and NS111416, and by start-up funds from Cornell University.

## References

1. Kucharski AJ, Russell TW, Diamond C, Liu Y, Edmunds J, Funk S, et al. Early dynamics of transmission and control of COVID-19: a mathematical modelling study. The lancet infectious diseases. 2020.

2. Peng L, Yang W, Zhang D, Zhuge C, Hong L. Epidemic analysis of COVID-19 in China by dynamical modeling. arXiv preprint arXiv:200206563. 2020.

3. Peak CM, Kahn R, Grad YH, Childs LM, Li R, Lipsitch M, et al. Modeling the comparative impact of individual quarantine vs. active monitoring of contacts for the mitigation of COVID-19. medRxiv. 2020.

4. Jewell NP, Lewnard JA, Jewell BL. Predictive mathematical models of the COVID-19 pandemic: Underlying principles and value of projections. Jama. 2020;323(19):1893–4.

5. Holmdahl I, Buckee C. Wrong but useful—what covid-19 epidemiologic models can and cannot tell us. New England Journal of Medicine. 2020.

6. Gatto M, Bertuzzo E, Mari L, Miccoli S, Carraro L, Casagrandi R, et al. Spread and dynamics of the COVID-19 epidemic in Italy: Effects of emergency containment measures. Proceedings of the National Academy of Sciences. 2020;117(19):10484–91.

7. Vespignani A, Tian H, Dye C, Lloyd-Smith JO, Eggo RM, Shrestha M, et al. Modelling COVID-19. Nature Reviews Physics. 2020:1–3.

8. Wu JT, Leung K, Leung GM. Nowcasting and forecasting the potential domestic and international spread of the 2019-nCoV outbreak originating in Wuhan, China: a modelling study. The Lancet. 2020;395(10225):689–97.

9. Ferguson N, Laydon D, Nedjati Gilani G, Imai N, Ainslie K, Baguelin M, et al. Report 9: Impact of non-pharmaceutical interventions (NPIs) to reduce COVID19 mortality and healthcare demand. 2020.

10. Kissler SM, Tedijanto C, Lipsitch M, Grad Y. Social distancing strategies for curbing the COVID-19 epidemic. medRxiv. 2020.

11. Prem K, Liu Y, Russell TW, Kucharski AJ, Eggo RM, Davies N, et al. The effect of control strategies to reduce social mixing on outcomes of the COVID-19 epidemic in Wuhan, China: a modelling study. The Lancet Public Health. 2020.

12. Giordano G, Blanchini F, Bruno R, Colaneri P, Di Filippo A, Di Matteo A, et al. Modelling the COVID-19 epidemic and implementation of population-wide interventions in Italy. Nature medicine. 2020:1–6.

13. Morris DH, Rossine FW, Plotkin JB, Levin SA. Optimal, near-optimal, and robust epidemic control. arXiv preprint arXiv:200402209. 2020.

14. Block P, Hoffman M, Raabe IJ, Dowd JB, Rahal C, Kashyap R, et al. Social network-based distancing strategies to flatten the COVID-19 curve in a post-lockdown world. Nature Human Behaviour. 2020:1–9.

15. Chinazzi M, Davis JT, Ajelli M, Gioannini C, Litvinova M, Merler S, et al. The effect of travel restrictions on the spread of the 2019 novel coronavirus (COVID-19) outbreak. Science. 2020;368(6489):395–400.

16. Ngonghala CN, Iboi E, Eikenberry S, Scotch M, MacIntyre CR, Bonds MH, et al. Mathematical assessment of the impact of non-pharmaceutical interventions on curtailing the 2019 novel Coronavirus. Mathematical biosciences. 2020:108364.

17. Zhang J, Litvinova M, Liang Y, Wang Y, Wang W, Zhao S, et al. Changes in contact patterns shape the dynamics of the COVID-19 outbreak in China. Science. 2020.

18. Leclerc QJ, Fuller NM, Knight LE, Funk S, Knight GM, Group CC-W. What settings have been linked to SARS-CoV-2 transmission clusters? Wellcome Open Research. 2020;5(83):83.

19. Middleton J, Reintjes R, Lopes H. Meat plants—a new front line in the covid-19 pandemic. British Medical Journal Publishing Group; 2020.

20. Barnett ML, Grabowski DC, editors. Nursing homes are ground zero for COVID-19 pandemic. JAMA Health Forum; 2020: American Medical Association.

21. Klompas M. Coronavirus disease 2019 (COVID-19): protecting hospitals from the invisible. American College of Physicians; 2020.

22. Chu C-M, Cheng VC, Hung IF, Chan K-S, Tang BS, Tsang TH, et al. Viral load distribution in SARS outbreak. Emerging infectious diseases. 2005;11(12):1882.

23. Chu C-M, Poon LL, Cheng VC, Chan K-S, Hung IF, Wong MM, et al. Initial viral load and the outcomes of SARS. Cmaj. 2004;171(11):1349–52.

24. Imai M, Iwatsuki-Horimoto K, Hatta M, Loeber S, Halfmann PJ, Nakajima N, et al. Syrian hamsters as a small animal model for SARS-CoV-2 infection and countermeasure development. Proceedings of the National Academy of Sciences. 2020;117(28):16587–95.

25. Golden J, Cline C, Zeng X, Garrison A, Carey B, Mucker E, et al. Human angiotensin-converting enzyme 2 transgenic mice infected with SARS-CoV-2 develop severe and fatal respiratory disease. bioRxiv. 2020.

26. Gandhi M, Rutherford GW. Facial Masking for Covid-19—Potential for “Variolation” as We Await a Vaccine. New England Journal of Medicine. 2020.

27. Lee DF, Drouin R, Pitsikas P, Rainbow AJ. Detection of an involvement of the human mismatch repair genes hMLH1 and hMSH2 in nucleotide excision repair is dependent on UVC fluence to cells. Cancer research. 2004;64(11):3865–70. PubMed PMID: 15172995.

28. Liu Y, Yan L-M, Wan L, Xiang T-X, Le A, Liu J-M, et al. Viral dynamics in mild and severe cases of COVID-19. The Lancet Infectious Diseases. 2020.

29. Gandhi M, Yokoe DS, Havlir DV. Asymptomatic transmission, the Achilles’ heel of current strategies to control COVID-19. Mass Medical Soc; 2020.

30. Kermack WO, McKendrick AG. Contributions to the mathematical theory of epidemics – I. Proc R Soc Med. 1927;115A:700–21.

31. Anderson RM, May RM. Infectious diseases of humans. Oxofrd, England: Oxford University Press; 1991.

32. Flacco ME, Martellucci CA, Bravi F, Parruti G, Mascitelli A, Mantovani L, et al. SARS-CoV-2 lethality decreased over time in two Italian Provinces. medRxiv. 2020.

33. The infection fatality rate of COVID-19 in Stockholm – Technical report, Public Health Agency of Sweden. 2020.

34. Harrington C, Ross L, Chapman S, Halifax E, Spurlock B, Bakerjian D. Nurse staffing and coronavirus infections in California nursing homes. Policy, Politics, & Nursing Practice. 2020;21(3):174–86.

35. Lloyd-Smith JO, Schreiber SJ, Kopp PE, Getz WM. Superspreading and the effect of individual variation on disease emergence. Nature. 2005;438(7066):355–9.

36. Lau MS, Grenfell B, Nelson K, Lopman B. Characterizing super-spreading events and age-specific infectivity of COVID-19 transmission in Georgia, USA. medRxiv. 2020.

37. Nacoti M, Ciocca A, Giupponi A, Brambillasca P, Lussana F, Pisano M, et al. At the epicenter of the Covid-19 pandemic and humanitarian crises in Italy: changing perspectives on preparation and mitigation. NEJM Catalyst Innovations in Care Delivery. 2020;1(2).

38. Cohan A. Brigham and Women’s nurses call for stronger safety measures after COVID cluster expands. Boston Herald. 2020.

39. State Data and Policy Actions to Address Coronavirus. https://www.kff.org/health-costs/issue-brief/state-data-and-policy-actions-to-address-coronavirus/: 2020.

40. Pandemic Experience in the Long-Term Care Sector. https://www.cihi.ca/sites/default/files/document/covid-19-rapid-response-long-term-care-snapshot-en.pdf?emktg_lang=en&emktg_order=1: 2020.

41. Fisman D, Lapointe-Shaw L, Bogoch I, McCready J, Tuite A. Failing our most vulnerable: COVID-19 and long-term care facilities in Ontario. medRxiv. 2020.

42. COVID-19 in Racial and Ethnic Minority Groups https://www.cdc.gov/coronavirus/2019-ncov/need-extra-precautions/racial-ethnic-minorities.html2020. Available from: https://www.cdc.gov/coronavirus/2019-ncov/need-extra-precautions/racial-ethnic-minorities.html.

43. Park SW, Sun K, Viboud C, Grenfell BT, Dushoff J. Potential roles of social distancing in mitigating the spread of coronavirus disease 2019 (COVID-19) in South Korea. medRxiv. 2020.

44. Shim E, Tariq A, Choi W, Lee Y, Chowell G. Transmission potential and severity of COVID-19 in South Korea. International Journal of Infectious Diseases. 2020.

