## Supplementary Materials for "Role of high-dose exposure in transmission hot zones as a driver of SARS-CoV2 dynamics"

### Supplementary Information

#### Contents

**1 Model formulation and calculating $R_0$** **1**

**2 Four extreme cases of different epidemic dynamics** **6**

**3 Condition (IIa)** **8**

**4 Model extensions** **9**

#### 1 Model formulation and calculating $R_0$

Let us demote the number of suscepibles by $x(t)$, the number of mildly and severely infected individuals by $y_1(t)$ and $y_2(t)$, respectively, and the amount of virus in the community and in the hot zones by $C(t)$ and $H(t)$, respectively. Then the model studied here can be described by the system of ODEs:

$$\dot{x} = -x[(\beta_{1H}+\beta_{2H})H+(\beta_{1C}+\beta_{2C})C], \tag{1}$$
$$\dot{y}_1 = x(\beta_{1C}C+\beta_{1H}H)-\gamma_1 y_1, \tag{2}$$
$$\dot{y}_2 = x(\beta_{2C}C+\beta_{2H}H)-\gamma_2 y_2, \tag{3}$$
$$\dot{z} = \gamma_1 y_1+\gamma_2 y_2, \tag{4}$$
$$\dot{C} = b_C y_1+B_C y_2-\alpha_C C, \tag{5}$$
$$\dot{H} = b_H y_1+B_H y_2-\alpha_H H, \tag{6}$$

with the initial condition

$$x(0)=x_0, \quad y_1(0)=y_{10}, \quad y_2(0)=y_{20}, \quad c(0)=c_0, \quad h(0)=h_0.$$

Here, we have $x+y_1+y_2+z=1$ (we choose this normalization for simplicity). If we express the values $C(t)$ and $H(t)$ from the last two equations in quasi steady state, we get

$$C = \frac{b_C y_1 + B_C y_2}{\alpha_C}, \quad H = \frac{b_H y_1 + B_H y_2}{\alpha_H},$$

and have the following system of three ODEs,

$$\begin{aligned} \dot{x} &= -[(a_{11} + a_{21})y_1 + (a_{12} + a_{22})y_2]x, &(7)\\ \dot{y}_1 &= x(a_{11}y_1 + a_{12}y_2) - \gamma_1 y_1, &(8)\\ \dot{y}_2 &= x(a_{21}y_1 + a_{22}y_2) - \gamma_2 y_2, &(9)\end{aligned}$$

where we introduced the following notations:

$$\begin{aligned} a_{11} &= \frac{\beta_{1C} b_C}{\alpha_C} + \frac{\beta_{1H} b_H}{\alpha_H}, \quad a_{12} = \frac{\beta_{1C} B_C}{\alpha_C} + \frac{\beta_{1H} B_H}{\alpha_H}, &(10)\\ a_{21} &= \frac{\beta_{2C} b_C}{\alpha_C} + \frac{\beta_{2H} b_H}{\alpha_H}, \quad a_{22} = \frac{\beta_{2C} B_C}{\alpha_C} + \frac{\beta_{2H} B_H}{\alpha_H}. &(11)\end{aligned}$$

To calculate the parameter $R_0$, we will use the next generation method (see [1], [2], and study the initial dynamics of system (7-9) where we start from small values of $y_1(0)$ and $y_2(0)$. Then $x \approx x_0$ and we can linearize equations (8-9), obtaining a linear system of ODEs for variables $(y_1, y_2)$ with the matrix

$$\begin{pmatrix} x_0 a_{11} - \gamma_1 & x_0 a_{12} \\ x_0 a_{21} & x_0 a_{22} - \gamma_2 \end{pmatrix}. \qquad (12)$$

The solution is given by

$$\begin{pmatrix} y_1 \\ y_2 \end{pmatrix} = C_1 \begin{pmatrix} v_1 \\ 1 \end{pmatrix} e^{\lambda_1 t} + C_2 \begin{pmatrix} u_1 \\ 1 \end{pmatrix} e^{\lambda_2 t}, \qquad (13)$$

where $\lambda_1$ and $\lambda_2$ are the eigenvalues of matrix (12), $(v_1, 1)^T$ and $(u_1, 1)^T$ are the corresponding eigenvectors and $C_1$ and $C_2$ are constants. The solution will increase initially if at least one of the eigenvalues has a positive real part. The characteristic polynomial for matrix $M$ is given by

$$\lambda^2 + c_1 \lambda + c_0,$$

where

$$c_0 = \gamma_1 + \gamma_2 - X_0(a_{11} + a_{22}), \quad c_1 = X_0^2(a_{11}a_{22} - a_{12}a_{21}) - X_0(a_{11}\gamma_2 + a_{22}\gamma_1) + \gamma_1\gamma_2.$$

To ensure that at least one of the two eigenvalues has a positive real part, we need to require

$$c_0 < 0 \quad \text{or} \tag{14}$$
$$c_1 < 0. \tag{15}$$

Before we proceed, let us make a convenient notation:

$$A_{ij} = \frac{a_{ij}}{\gamma_i}, \quad i, j \in \{1, 2\},$$

that is,

$$A_{11} = \frac{1}{\gamma_1}\left(\frac{\beta_{1C}b_C}{\alpha_C} + \frac{\beta_{1H}b_H}{\alpha_H}\right), \quad A_{12} = \frac{1}{\gamma_1}\left(\frac{\beta_{1C}B_C}{\alpha_C} + \frac{\beta_{1H}B_H}{\alpha_H}\right), \tag{16}$$
$$A_{21} = \frac{1}{\gamma_2}\left(\frac{\beta_{2C}b_C}{\alpha_C} + \frac{\beta_{2H}b_H}{\alpha_H}\right), \quad A_{22} = \frac{1}{\gamma_2}\left(\frac{\beta_{2C}B_C}{\alpha_C} + \frac{\beta_{2H}B_H}{\alpha_H}\right). \tag{17}$$

If we assume that being infected through $C$ only results in mild infections, and being infected through $H$ only results in severe infections, we have the following simplified expressions, see the schematic in figure S1:

$$A_{11} = \frac{\beta_{1C}b_C}{\gamma_1\alpha_C}, \quad A_{12} = \frac{\beta_{1C}B_C}{\gamma_1\alpha_C}, \quad A_{21} = \frac{\beta_{2H}b_H}{\gamma_2\alpha_H}, \quad A_{22} = \frac{\beta_{2H}B_H}{\gamma_2\alpha_H}. \tag{18}$$

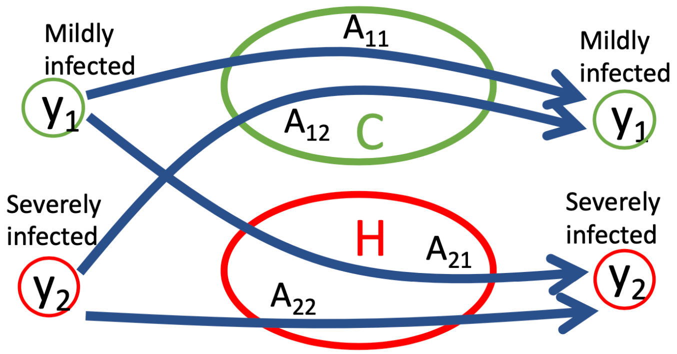


Figure S1: A schematic showing the meaning of the four matrix elements in the case where infection through C (H) results in only mild (severe) infections.

We have

$$c_1 = \gamma_1\gamma_2\left(X_0^2 Det - X_0 Tr + 1\right),$$

where $Det$ and $Tr$ are the determinant and the trace of the matrix

$$\begin{pmatrix} A_{11} & A_{12} \\ A_{21} & A_{22} \end{pmatrix},$$

respectiely. Let us further denote

$$A_{min} = \min\{A_{11}, A_{22}\}, \quad A_{max} = \max\{A_{11}, A_{22}\}.$$

Condition (14) gives

$$X_0 > \frac{\gamma_1 + \gamma_2}{A_{11}\gamma_1 + A_{22}\gamma_2} \equiv \hat{X}_0,$$

where the following inequality holds:

$$\frac{1}{A_{max}} \le \hat{X}_0 \le \frac{1}{A_{min}}. \tag{19}$$

Next, we consider condition (15). Denote by $\hat{X}_1$ and $\hat{x}_2$ the roots of $c_1$:

$$\hat{X}_1 = \frac{Tr - \sqrt{Tr^2 - 4Det}}{2Det}, \quad \hat{X}_2 = \frac{Tr + \sqrt{Tr^2 - 4Det}}{2Det}.$$

We have the following two cases.

- If $Det > 0$, we have $0 \le \hat{X}_1 \le \hat{X}_2$, and condition (15) is equivalent to $\hat{X}_1 < X_0 < \hat{X}_2$. It is possible to show that $\hat{X}_1 < \frac{1}{A} < \hat{X}_2$, where $A$ can be both $A_{11}$ and $A_{22}$ (see below). Therefore, we have $\hat{X}_1 < \hat{X}_0 < \hat{X}_2$, and condition(14,15) is equivalent to $X_0 > \hat{X}_1$.
- If $Det < 0$, we can see that $\hat{X}_2 < 0 < \hat{X}_1$, and condition (15) is equivalent to $X_0 < \hat{X}_2$ or $X_0 > \hat{X}_1$. It is further possible to show that $\hat{X}_1 < \frac{1}{A}$, where $A$ can be both $A_{11}$ and $A_{22}$ (see below). Therefore, we have $\hat{X}_1 < \hat{X}_0$, and condition(14,15) is again equivalent to $X_0 > \hat{X}_1$.

Before we continue, let us prove the inequalities used in the above argument. If $Det > 0$ then $\hat{X}_1 < 1/A$ is equivalent to $A\sqrt{Tr^2 - 4Det} > ATr - 2Det$, and $\hat{X}_2 > 1/A$ is equivalent to $A\sqrt{Tr^2 - 4Det} > 2Det - ATr$. For both of these inequalities, the left hand side is positive. When squared, they result in the following inequality:

$$Det[(A - A_{22})(A - A_{11}) - A_{12}A_{21}] < 0, \tag{20}$$

which is true if $A$ is either $A_{11}$ or $A_{22}$. This implies that the double inequality $\hat{X}_1 < \frac{1}{A} < \hat{X}_2$ holds.

Similarly, if $Det < 0$ then $\hat{X}_1 < 1/A$ is equivalent to $A\sqrt{Tr^2 - 4Det} < ATr - 2Det$, which when squared leads to the opposite of inequality (20), which (under $Det < 0$) holds if either $A = A_{11}$ or $A = A_{22}$.

We conclude that an epidemic will rise from low initial numbers of infecteds if $X_0 > \hat{X}_1$, which in terms of the original system translates into $x_0 > \hat{X}_1$, and the condition for successful epidemic is equivalent to

$$R_0 \equiv \frac{x_0}{\hat{X}_1} > 1,$$

where $x_0$ is the initial population of susceptibles.

A convenient way to write $R_0$ is as follows:

$$R_0 = x_0 r, \quad r = \frac{2(A_{11}A_{22} - A_{12}A_{21})}{A_{11} + A_{22} - \sqrt{(A_{11} - A_{22})^2 + 4A_{12}A_{21}}}, \tag{21}$$

There are several particular cases of interest.

- If $A_{12} = 0$ or $A_{21} = 0$ (the absence of at least one of the cross-infection pathways), we have $Det > 0$, $\hat{X}_1 = 1/A_{max}$, $\hat{X}_2 = 1/A_{min}$, and we have

  $$R_0 = x_0 A_{max}.$$

  If $A_{22} = A_{max}$ we have

  $$R_0 = x_0 \left( \frac{\beta_{2C} B_C}{\gamma_2 \alpha_C} + \frac{\beta_{2H} B_H}{\gamma_2 \alpha_H} \right)$$

- If $A_{11} \approx A_{22}$ (more precisely, $|A_{11} - A_{22}| \ll 2\sqrt{A_{12}A_{21}}$), we obtain

  $$R_0 = x_0(A_{11} + \sqrt{A_{12}A_{21}}).$$

  If the two types are identical, we have $A_{21} = A_{12}$ and

  $$R_0 = x_0(A_{11} + A_{12}).$$

  For identical types without cross-infection, we have

  $$R_0 = x_0 A_{11}.$$

- If only one type is present, then $x_0$ must exceed the expression corresponding to the existing type, that is, $R_0 = x_0 A_{ii}$ if only type $i$ is present.

- If $Det = 0$, then $R_0 = x_0 Tr$, that is,

$$R_0 = x_0(A_{11} + A_{22}).$$

### 2 Four extreme cases of different epidemic dynamics

Let us denote the infection and deposition rates normalized by the rate of removal as follows:

$$\tilde{b}_C = \frac{b_C}{\alpha_C}, \quad \tilde{b}_H = \frac{b_H}{\alpha_H}, \quad \tilde{B}_C = \frac{B_C}{\alpha_C}, \quad \tilde{B}_H = \frac{B_H}{\alpha_H}, \tag{22}$$

$$\tilde{\beta}_{1C} = \frac{\beta_{1C}}{\gamma_1}, \quad \tilde{\beta}_{1H} = \frac{\beta_{1H}}{\gamma_1}, \quad \tilde{\beta}_{2C} = \frac{\beta_{2C}}{\gamma_2}, \quad \tilde{\beta}_{2H} = \frac{\beta_{2H}}{\gamma_2}. \tag{23}$$

We can express these quantities as matrix elements:

$$Q_{\text{acq}} = \begin{pmatrix} \tilde{\beta}_{1C} & \tilde{\beta}_{1H} \\ \tilde{\beta}_{2C} & \tilde{\beta}_{2H} \end{pmatrix}, \quad Q_{\text{dep}} = \begin{pmatrix} \tilde{b}_C & \tilde{B}_C \\ \tilde{b}_H & \tilde{B}_H \end{pmatrix} \tag{24}$$

where the subscripts "acq" and "dep" stand for "acquisition" and "deposition" of infection, and the two matrices describe the process of getting infected from C or H, and the process of virus deposition into pools C and H, respectively. Then, the matrix elemets $A_{ij}$ are given by the following matrix product:

$$\{A_{ij}\} = Q_{\text{acq}} Q_{\text{dep}}. \tag{25}$$

It is easy to show that the value of $r$ in expression (21) is the larger of the eigenvalues of matrix $\{A_{ij}\}$, that is, the larger of the solutions of

$$(r - A_{11})(r - A_{22}) = A_{12}A_{21},$$

given by

$$r = \frac{1}{2}(TrA + \sqrt{TrA^2 - 4DetA}).$$

| Parameter | $\gamma_1$ | $\gamma_2$ | $\alpha_C$ | $\alpha_H$ | $\beta_{1C}$ | $\beta_{1H}$ | $\beta_{2C}$ | $\beta_{2H}$ | $b_C$ | $b_H$ | $B_C$ | $B_H$ |
|---|---|---|---|---|---|---|---|---|---|---|---|---|
| During | $1/F_C$ | $1/F_H$ | $1/F_C$ | $1/F_H$ | $F_C$ | $F_H$ | $F_C$ | $F_H$ | $F_C$ | $F_H$ | $F_C$ | $F_H$ |
| After | 1 | $1/F_H$ | 1 | $1/F_H$ | 1 | $F_H$ | 1 | $F_H$ | 1 | $F_H$ | 1 | $F_H$ |

Table 1: Model parameters and the multiplicative factors that modify them during and after the social distancing phase.

Of interest is to determine the condition that specifies which population, $y_1$ or $y_2$, dominates the infected individuals. To determine whether $y_1$ or $y_2$ is larger, consider solution (13) and assume that $\lambda_1 > \lambda_2$. Then the population of mildly infected individuals is larger than that of severely infected individuals if $v_1 > 1$. We have

$$v_1 = \frac{q + \sqrt{q^2 + 4a_{12}a_{21}}}{2a_{21}}, \quad \text{where } q = x_0(a_{11} - a_{22}) - \gamma_1 + \gamma_2,$$

and $v_1 > 1$ is equivalent to

$$x_0(a_{11} + a_{12}) - \gamma_1 > x_0(a_{22} + a_{21}) - \gamma_2. \tag{26}$$

If we rewrite this in terms of $A_{ij}$ we obtain the condition

$$\gamma_1 A_{12} - \gamma_2 A_{21} > \gamma_2(A_{22} - 1) - \gamma_1(A_{11} - 1).$$

In the special case where $\gamma_1 = \gamma_2$ we obtain the condition

$$A_{11} + A_{12} > A_{21} + A_{22}.$$

Depending on the parameters, there are 4 types of extreme cases, which differ by the following features:

- The main contribution to infection spread could be (1) by mildly infected, $A_{11} > A_{22}$ or (2) by severely infected patients, $A_{11} < A_{22}$.
- The majority of infected individuals could be (a) mildly infected (condition (26)) or (b) severely infected (the opposite condition).

There are 4 combinations: (Ia), (Ib), (IIa), and (IIb). Cases I(a) is illustrated in main figure 2(a) and case II(a)) in the main figure 2(b-d). The other two

| Figure | $t_1$ | $t_2$ | $F_C$ | $F_H$ |
|---|---|---|---|---|
| Main 2(a) | 50 | 100 | 0.5 | 0.7 |
| Main 2(c) | 55 | 100 | 0.3 | 0.87 |
| Main 2(d) | 65 | 100 | 0.5 | 0.7 |
| Main 2(e) | 50 | 100 | 0.5 | 0.8 |
| Main 2(f) | 50 | 100 | 0.5 | 0.5 |
| S2(a) | 50 | 100 | 0.5 | 0.7 |
| S2(b) | 70 | 100 | 0.5 | 0.7 |

Table 2: Intervention parameters used in the simulations. Here social distancing starts at time $t_1$ and is relaxed at time $t_2$, and the parameters during and after the distancing phase are modified according to the scheme in table 1.

cases, (I(b) and II(b)), can be found in figure S2(a,b). The parameter values for all four cases illustrated are summarized in panel (c) of figure S2.

Both in main figure 2 and in figure S2(a,b), we assume that during social distancing (dashed lines), and afterwards, the parameter values are modified in the way described in table 1. For the simulations that are presented, the social distancing parameters are listed in table 2.

### 3 Condition (IIa)

Of particular interest is case (IIa). For this to hold, in the particular case whee $\gamma_1 = \gamma_2$, we must have

$$A_{22} > A_{11}, \quad A_{12} > A_{21},$$

which is equivalent to the inequality

$$\frac{b_H}{B_C} < \frac{\beta_1}{\beta_2} < \frac{B_H}{b_C},$$

which requires a simple necessary condition

$$B_C B_H > b_C b_H.$$

In figure S3 we studied the sensitivity of the value of $R$ to parameter values, by varying them one by one around fixed values. We can see that

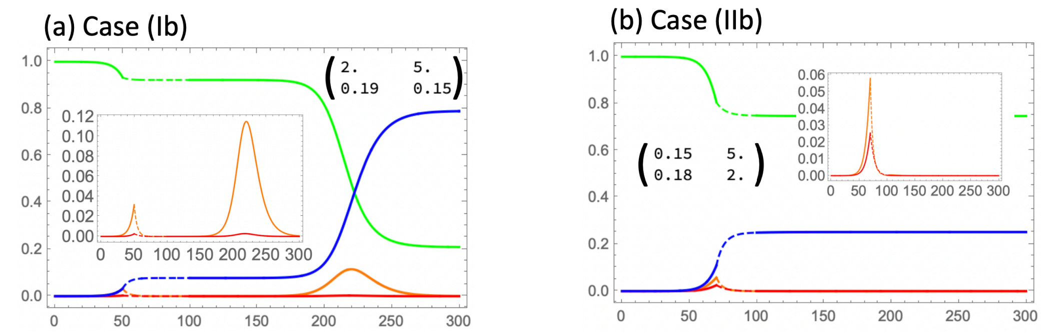


(c) Parameter values

| Case | Figure | $\gamma_1$ | $\gamma_2$ | $\alpha_C$ | $\alpha_H$ | $\beta_{1C}$ | $\beta_{1H}$ | $\beta_{2C}$ | $\beta_{2H}$ | $b_C$ | $b_H$ | $B_C$ | $B_H$ |
|---|---|---|---|---|---|---|---|---|---|---|---|---|---|
| (Ia) | Main 2(a) | 0.14 | 0.25 | 2. | 0.33 | 0.01 | 0.0005 | 0.001 | 0.005 | 60. | 1.6 | 85. | 1.3 |
| (Ib) | S2(a) | 0.14 | 0.25 | 2. | 0.33 | 0.1 | 0.0005 | 0.001 | 0.005 | 5.6 | 3. | 14. | 2. |
| (IIa) | Main 2(b-d) | 0.14 | 0.25 | 2. | 0.33 | 0.01 | 0.0005 | 0.001 | 0.005 | 3.4 | 3. | 130. | 29. |
| (IIb) | S2(b) | 0.14 | 0.25 | 2. | 0.33 | 0.1 | 0.0005 | 0.001 | 0.005 | 0.34 | 3. | 13. | 33. |

Figure S2: Extreme cases and simulation parameters. Examples of cases (Ib) and (IIb) are presented in panels (a) amd (b), respectively. The populations are shown as functions of time: susceptibles (green), mildly infected (orange), severely infected (red), and recovered/dead (blue). The period of social distancing is shown by dashed lines. The matrix $\{A_{ij}\}$ is presented in parentheses. The dynamics of infecteds are also shown in the insets for greater detail. Parameter values for the examples illustrated here and also in main figure 2(a-d) are listed in panel (c).

changes in the parameters related to zone H have the largest effect on $R$ in this case. Not surprisingly, changes in these parameters regardless of relaxation in C-related parameters will keep the epidemic from spreading. Panel (c) shows the extent of region (IIa) in the space of infection deposition parameters, for a given set of fixed acquisition parameters.

### 4 Model extensions

In order to include more realism in the model, we first included a saturation term in the transmission mechanism, where parameter $\epsilon$ defines the level of saturation of the infection. We further included a mechanism where the probability of acquiring a severe infection through channel C or through channel H depends on the amount of infection: the higher the value of C and

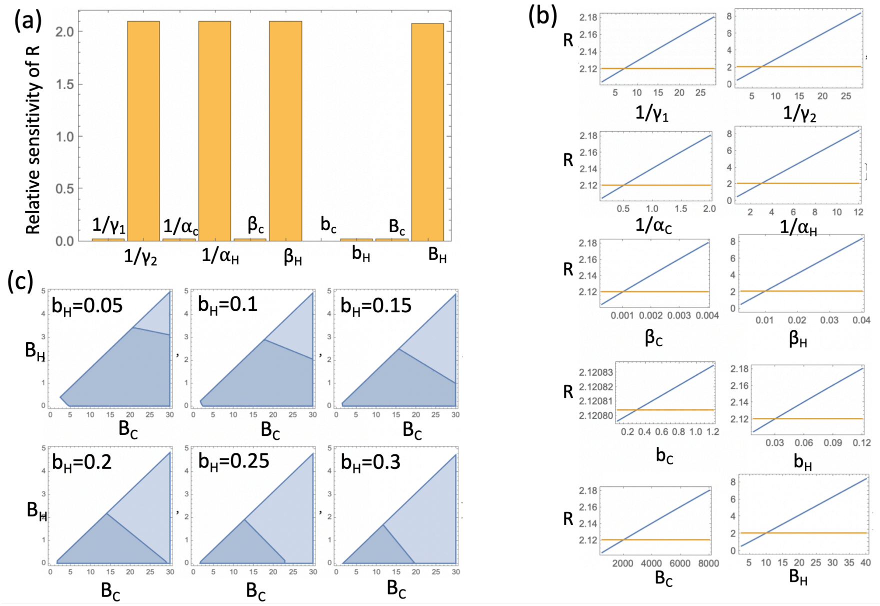


Figure S3: Case II(a). Sensitivity analysis of parameter $R$ (with $x = 1$) is presented in panels (a) and (b). The dependence of $R$ on individual parameters (which are varied between 25 % and 4000% of their chosen values) is shown in (b) (the horizontal line shows the value corresponding to the fixed parameters chosen). Panel (a) shows that relative sensitivity $q\frac{\partial R}{\partial q}$, where $q$ is each of the parameters. The parameters (unless they are varied) are set to the following values: $\gamma_1 = \gamma_2 = 1/7, \alpha_C = 2, \alpha_H = 1/3, \beta_{1C} = 0.001, \beta_{2H} = 0.01, \beta_{1H} = \beta_{2C} = 0, b_C = 0.3, b_H = 0.03, B_C = 2000, B_H = 10$. Panel (c) shows slices of a 3D parameters spaces (with varying parameters $B_H, B_C, b_H$ where the system belongs to regime 2(a) (blue) and $1 < R < 5$ (the darker regions). The rest of the parameters are: $\gamma_1 = 1/4, \gamma_2 = 1/4, \alpha_C = 2, \alpha_H = 1/3, \beta_{1C} = \beta_{2H} = 0.1, \beta_{1H} = \beta_{2C} = 0, b_C = 0.01$.

H, the more likely it is that the infection is severe. These mechanisms were incorporated in equations (2-3) in the following way:

$$\dot{y}_1 = x\left(\beta_{2C}\frac{\epsilon C}{C+\epsilon_C}\ \frac{C_C^\nu}{C_C^\nu + C^\nu} + \beta_{2H}\ \frac{\epsilon H}{H+\epsilon}\frac{H_c^\nu}{H_C^\nu + H^\nu}\right) - \gamma_1 y_1, \quad (27)$$

$$\dot{y}_2 = x\left(\beta_{2C}\frac{\epsilon C}{C+\epsilon_C}\ \frac{C^\nu}{C_C^\nu + C^\nu} + \beta_{2H}\ \frac{\epsilon_H H}{H+\epsilon_H}\frac{H^\nu}{H_C^\nu + H^\nu}\right) - \gamma_2 y_2. \quad (28)$$

Equation (1) was also modified accordingly. In these equations, saturation terms are included both in terms describing infection through channel $H$ and channel $C$. Therefore, we have two corresponding parameters, $\epsilon_H$ and $\epsilon_C$, that reflect at which level saturation occurs in each term. Further, we included Hill-type functions to modulate infection probabilities. For example, when infection happens through channel H, constant $H_C$ is the level of infection at which it becomes more likely to become severely infected than mildly infected through H channel, and power $\nu$ characterizes the steepness of the transition from "mostly mild" to "mostly severe". Similarly, for transmission through channel C, parameter $C_C$ reflects at which level of infection in the community most individuals will become severely infected. While these additional parameters are introduced, parameters $\beta_{1H}$ and $\beta_{2C}$ are eliminated, being replaced by the new terms in the equations for $y_1$ and $y_2$. Simulations of the resulting system are presented in main figure 3. Parameter values used in that figure are listed in figure S4.

| | $\gamma_1$ | $\gamma_2$ | $\alpha_C$ | $\alpha_H$ | $\beta_{1C}$ | $\beta_{1H}$ | $\beta_{2C}$ | $\beta_{2H}$ | $b_C$ | $b_H$ | $B_C$ | $B_H$ |
|---|---|---|---|---|---|---|---|---|---|---|---|---|
| Fig 2(e,f) | 0.14 | 0.25 | 2. | 0.33 | 0.01 | 0.0005 | 0.001 | 0.005 | 37. | 1.8 | 220. | 11. |
| Fig 3(a,b) | 0.071 | 0.14 | 2. | 0.33 | 0. | 0. | 0.1 | 0.1 | 3. | 0.3 | 500. | 50. |

Figure S4: Parameter values for main figures 2(e,f) and 3. In addition, intervention parameters for main figure 2(e,f) are given in table 2. For main figure 3, $H_C = 0.2, C_C = 10, \nu = 0.5, \epsilon_C = \epsilon_C = 10$.

### References


[1] Odo Diekmann, Johan Andre Peter Heesterbeek, and Johan AJ Metz. On the definition and the computation of the basic reproduction ratio R0 in models for infectious diseases in heterogeneous populations. *Journal of mathematical biology*, 28(4):365–382, 1990.

[2] Pauline Van den Driessche and James Watmough. Reproduction numbers and sub-threshold endemic equilibria for compartmental models of disease transmission. *Mathematical biosciences*, 180(1-2):29–48, 2002.
